# Ethnic homophily affects vaccine prioritization strategies

**DOI:** 10.1101/2022.07.15.22277696

**Authors:** Claus Kadelka, Md Rafiul Islam, Audrey McCombs, Jake Alston, Noah Morton

**Affiliations:** Department of Mathematics, Iowa State University, Ames, IA 50011, USA; Department of Statistics, Iowa State University, Ames, IA 50011, USA

**Keywords:** COVID-19, compartmental disease model, vaccine roll-out, homophily, ethnicity

## Abstract

People are more likely to interact with other people of their ethnicity—a phenomenon known as ethnic homophily. In the United States, people of color are known to hold proportionately more high-contact jobs and are thus more at risk of virus infection. At the same time, these ethnic groups are on average younger than the rest of the population. This gives rise to interesting disease dynamics and non-trivial trade-offs that should be taken into consideration when developing prioritization strategies for future mass vaccine roll-outs.

Here, we study the spread of COVID-19 through the U.S. population, stratified by age, ethnicity, and occupation, using a detailed, previously-developed compartmental disease model. Based on historic data from the U.S. mass COVID-19 vaccine roll-out that began in December 2020, we show, (i) how ethnic homophily affects the choice of optimal vaccine allocation strategy, (ii) that, notwithstanding potential ethical concerns, differentiating by ethnicity in these strategies can improve outcomes (e.g., fewer deaths), and (iii) that the most likely social context in the United States is very different from the standard assumptions made by models which do not account for ethnicity and this difference affects which allocation strategy is optimal.

**Highlights:**

- A social mixing model accounting for ethnic homophily and variable job-related risk level is developed.
- A scenario that differs strongly from standard homogeneous mixing assumptions best matches U.S. ethnicity-specific death and case counts.
- Two trade-offs are explored: Should (i) old or young, and (ii) people of color or White and Asian people first receive COVID-19 vaccines?
- Exhaustive simulation of a compartmental disease model identifies the optimal allocation strategy for different demographic groups.
- Optimal strategies depend on the underlying mixing pattern and strategies that differentiate vaccine access by ethnicity outperform others.

## 1. Introduction

In the United States, people of color (POC) have been disproportionately affected by COVID-19 (CDC, 2020). Overall, POC (people from the following racial/ethnic backgrounds: Hispanic/Latino, American Indian/Alaskan Native, Black, Native Hawaiian/Other Pacific Islander, and mixed-race/ethnicity) comprise 34.1% of the U.S. population but suffered 42.2% of COVID-19 cases, while non-Hispanic White and non-Hispanic Asian (WA) make up 65.9% of the population but comprised 57.8% of total cases (Kaiser Family Foundation, 2022). Studies from earlier in the pandemic report even greater disparities. Adjusting for age and other relevant factors, the infection rate in predominantly Black counties in the U.S. in 2020 was over three times that of predominantly white counties, and the Navajo Nation had more cases per capita than any state in the country (Shadmi et al., 2020). In the city of Chicago, over 50% of COVID-19 deaths were among the Black population, which only comprises about 30% of the city population (Bhala et al., 2020).

New York City reported approximately twice as many deaths per capita in the Black and Latino populations than in the White population (Hooper et al., 2020). And overall, across the U.S. the 20% of disproportionately Black counties accounted for 52% of cases and 58% of COVID-19 deaths nationwide (Millett et al., 2020).

The reasons for these disparities are complex, but one common explanation is that POC tend to live in more crowded conditions, and are more likely to work in public-facing, high-contact occupations, which frequently do not allow for physical distancing (e.g., services and transportation) (Hooper et al., 2020; Bhala et al., 2020). Homophily describes the tendency of people from a particular demographic group to interact more frequently with people from the same group, and homophily among ethnic groups as well as age groups is well documented (McPherson et al., 2001; Currarini et al., 2009; Mollica et al., 2003). Homophily is known to affect disease dynamics (Kadelka and McCombs, 2021; Burgio et al., 2022; Hiraoka et al., 2022; Salathé and Bonhoeffer, 2008), and while the 4-phase COVID-19 vaccine allocation strategy implemented by the CDC in 2020 accounted for some differences in occupational hazards (healthcare workers first, then frontline essential workers, etc.), it did not account for ethnic homophily (Dooling et al., 2020).

COVID-19 also has disproportionate effects on older people. Only 16.5% of the U.S. population is over the age of 65, but 75.1% of deaths due to COVID-19 were in this age group. The fundamental trade-off when aiming to minimize COVID-19 deaths through an optimal vaccine roll-out therefore concerns age: Should the older population be vaccinated first to directly reduce mortality, or should younger people be vaccinated first because they have more contacts (Mossong et al., 2008; Prem et al., 2017), thereby reducing the spread of the disease and indirectly reducing mortality? Taking ethnicity into account further complicates the answer to this question. The WA population is demographically older than POC: 20.2% of WA are over age 65 while only 9.3% of POC fall into this age group (US Census Bureau, 2020). Complex interactions between demographics and contact rates may therefore result in an optimal allocation strategy with counter-intuitive prioritizations of both age groups and ethnicity groups.

The CDC COVID-19 vaccine allocation strategy assigned vaccine access based on age and occupational hazards but not based on ethnicity (Dooling et al., 2020). Most subsequent research into optimal vaccination strategies also primarily considered the fundamental vaccine prioritization trade-off concerning age (Bubar et al., 2021; Matrajt et al., 2021b,a; Foy et al., 2021). Some studies accounted for occupation as well as known COVID-19 risk factors, in addition to age (Islam et al., 2021; März et al., 2022), but to our knowledge, only one study incorporates ethnicity, as a predictor in a simple logistic regression model (McDonald et al., 2020).

In this study, we build on our previously-developed compartmental disease model, which stratified the U.S. population by age, living conditions, comorbidities, and occupation and was designed to accurately evaluate the 2020 CDC vaccine allocation strategy (Islam et al., 2021). Here we expand the model by incorporating differential contact rates and occupational hazards by age and ethnicity group to investigate how the fundamental trade-off and the trade-off regarding ethnicity might affect the optimal vaccine allocation (Figure 1). We separate the population into ten groups stratified by age (4 age classes), ethnic group (POC and WA), and occupational risk level (high and low risk) and implement a new method for constructing contact matrices with specified levels of ethnic homophily and social interaction (Kadelka, 2022). A global optimization algorithm identifies the optimal strategy from all 2.9 million possibly optimal allocation strategies which structure the vaccine roll-out in 1-5 or 10 phases. We investigate, (i) how ethnic homophily and social interaction parameters affect the choice of optimal vaccine allocation strategy, and (ii) notwithstanding possible ethical concerns, whether differentiating by ethnicity in these strategies can lead to better societal outcomes (e.g., fewer deaths).

**Figure 1:**
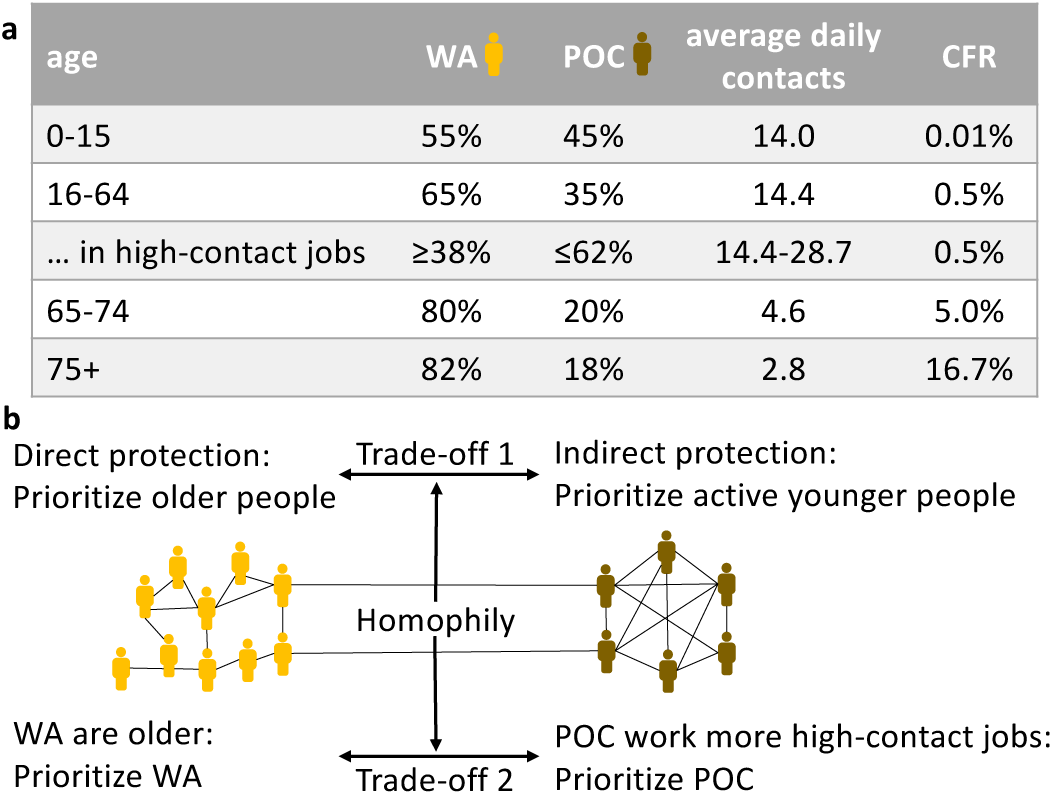
Trade-offs in vaccination prioritizations. (a) For each age group, the ethnic decomposition (into non-Hispanic White or Asian, WA versus Hispanic or people of color, POC) in the United States, as well as average daily contacts and case fatality rate (CFR) are shown. For the working-age population (16-64 year olds) employed in high-contact jobs, ranges for the ethnic decomposition and the average daily contact numbers, two key model parameters, are shown. (b) Infografic highlighting the two fundamental vaccine allocation trade-offs investigated in this study, which both depend on the specific contact structure of the U.S. population including the level of ethnic homophily.

## 2. Methods

### 2.1. Model overview

We study the spread of SARS-CoV-2 by adapting a previously published, detailed compartmental disease model (Islam et al., 2021) used to evaluate the U.S. COVID-19 vaccine allocation strategy. To ensure an accurate evaluation, this model accounts for key virus, disease, social and behavioral elements of the COVID-19 pandemic (e.g., age-dependent susceptibility to infection and case fatality rate, population-wide social distancing levels that depend on the current number of active cases, exact historic vaccination rates, and emergence of virus variants with higher transmissibility). For details, see Islam et al. (2021).

In this study, we split the entire U.S. population into ten sub-populations based on age (0 − 15, 16 − 64, 65 − 74, and 75+ years of age), as well as ethnicity and type of occupation for the working-age population of 16 − 64 year olds (Figure 1a). We consider only a binary stratification with respect to ethnicity by combining White and Asian into one group (WA) and all other ethnicities disproportionately affected by COVID-19 into the other group (POC) (Kaiser Family Foundation, 2022; Shadmi et al., 2020; Khanijahani et al., 2021). According to the 2020 U.S. census (US Census Bureau, 2020), there are 64.8 million children under the age of 16, 209.4 million 16 − 64 year olds, 31.5 million 65 − 74 year olds, and 22.6 million 75 and older. The proportion of people reported as WA among each of these four age groups is 55.0%, 65.4%, 79.6%, and 82.3%, respectively.

We also consider a binary stratification for occupation. We distinguish between *low-contact* (LC) jobs, which can be performed virtually and/or require few close human contacts (e.g., most office jobs), and *high-contact* (HC) jobs (e.g., most jobs in health care, meat packing plants, factories). People of color are known to work proportionately more high-contact jobs (Tai et al., 2021). While the U.S. Bureau of Labor provides a distribution based on ethnicity for different job categories (US Bureau of Labor Statistics, 2021a,b), the reported job categories do not break cleanly into LC and HC jobs, so it is not clear how this can accurately inform the stratification of ethnicity across job types. In the absence of accurate data, we assume that 25% of all 16-64 year olds are employed in HC jobs. Further, we define the ratio of the proportion of POC versus the proportion of WA employed in high-contact jobs as one of the three key model parameters we vary in this study, denoted *ψ*. That is,

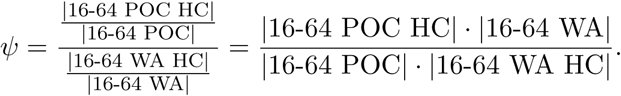

We consider *ψ* ∈ {1, 3} where *ψ* = 3 means POC are three times more likely to work high-contact jobs. Note that all 16-64 year old individuals who are not working are grouped together with those in low-contact jobs.

### 2.2. Contact patterns

The entry *x*_*i,j*_ in a square *contact matrix* describes the average number of daily interactions an individual in sub-population *i* has with individuals from sub-population *j*. Extensive, diary-based surveys in eight European countries have revealed empirical contact matrices stratified by age (Mossong et al., 2008). More recently, using surveys and demographic data, age-stratified contact matrices have been inferred for many countries, including the United States (Prem et al., 2017). For all countries, the contact matrices exhibit homophily with respect to age (also known as age-assortativity), meaning there are more contacts between people of similar age (i.e., values closer to the diagonal of the contact matrix are higher). We use here the age-stratified contact matrix for the United States.

To describe the level of ethnic homophily in contact patterns, let *p* ∈ (0, 1) be the proportion of WA individuals and let *φ* ∈ [0, 1] be the proportion of all contacts that occur between people of the same ethnicity group (WA or POC). We have 𝔼(*φ*) = *p*^2^ + (1 − *p*)^2^. As in Kadelka and McCombs (2021) and similar to Feng et al. (2015), we define ethnic homophily, denoted *h*, as the difference between the observed and expected proportions of all contacts that occur between people of different ethnicity group (WA and POC), and scale it so that *h* ∈ [−1, 1].

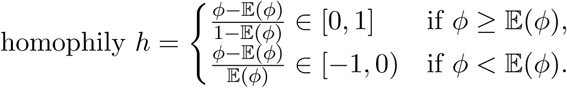

For completeness, this definition includes the case of heterophily, i.e., *φ <* 𝔼(*φ*). If *φ* = 1, we have 100% homophily, corresponding to complete segregation of WA and POC. The other extreme case of *φ* = 0 corresponds to 100% heterophily, in which case a graph describing all contacts would be bipartite. While the exact level of ethnic homophily is not known, several surveys indicate a high positive value in the range of 70 − 90% (Ingraham, 2014; Cox et al., 2016). In this study, we therefore only consider *h* ∈ [0, 1]. Albeit unrealistic, we include *h* = 0% (i.e., *φ* = 𝔼(*φ*)) as lower bound as it represents the intrinsic assumption used in any model that does not account for ethnic homophily.

A third key model parameter accounts for the increased level of average daily contacts due to employment in a high-contact job setting (compared to a low-contact job setting), denoted *κ*. We consider *κ* ∈ {1, 3}, where *κ* = 1 implies no difference in the contact patterns between people employed in high- and low-contact jobs, and *κ* = 3 means individuals in high-contact jobs have three times as many daily contacts compared to people in low-contact jobs. That is, for each *j* = 1, …, 10,

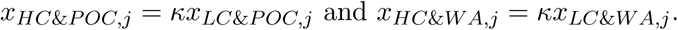

The binary classification of occupation and the lack of data on average contact levels per occupation make it hard to estimate a reasonable choice for *κ*. However, the true value for *κ* will most likely be somewhere within the considered range.

To obtain the contact matrix for our study, we start with a 4 × 4 age-stratified contact matrix inferred for the United States (Prem et al., 2017). Due to the undirected nature of physical contacts, any physical contact matrix relevant for the study of infectious diseases should be reciprocal (also known as symmetric) with respect to population size. That is, *n*_*i*_*x*_*i,j*_ = *n*_*j*_*x*_*j,i*_ for all *i, j* where *n*_*i*_ is the number of individuals in sub-population *i*. Using a standard procedure (Funk et al., 2019), we transform the empirical 4 × 4 age-based contact matrix, which for various reasons is typically quite far from reciprocal, into a reciprocal contact matrix. To then infer a 10 × 10 contact matrix that accurately describes the contact patterns between the ten sub-populations—including ethnic homophily and higher contact rates for individuals employed in high-contact jobs—we follow the recent procedure described in Kadelka (2022). The resulting contact matrix remains reciprocal and has the following four desirable properties.

1. It possesses a defined level of ethnic homophily.
2. WA and POC people of the same age and same occupation type have the same overall number of contacts.
3. An individual of age *i* has exactly the same average number of daily contacts with individuals of age *j* as defined in the reciprocal 4 × 4 contact matrix.
4. Individuals of a given ethnicity group with high-contact jobs have the exact same contact patterns as individuals of that ethnicity group with low-contact jobs, except they have *κ* ≥ 1 more contacts.

Figure 2 shows the contact matrix for some of the different scenarios considered in this study.

**Figure 2:**
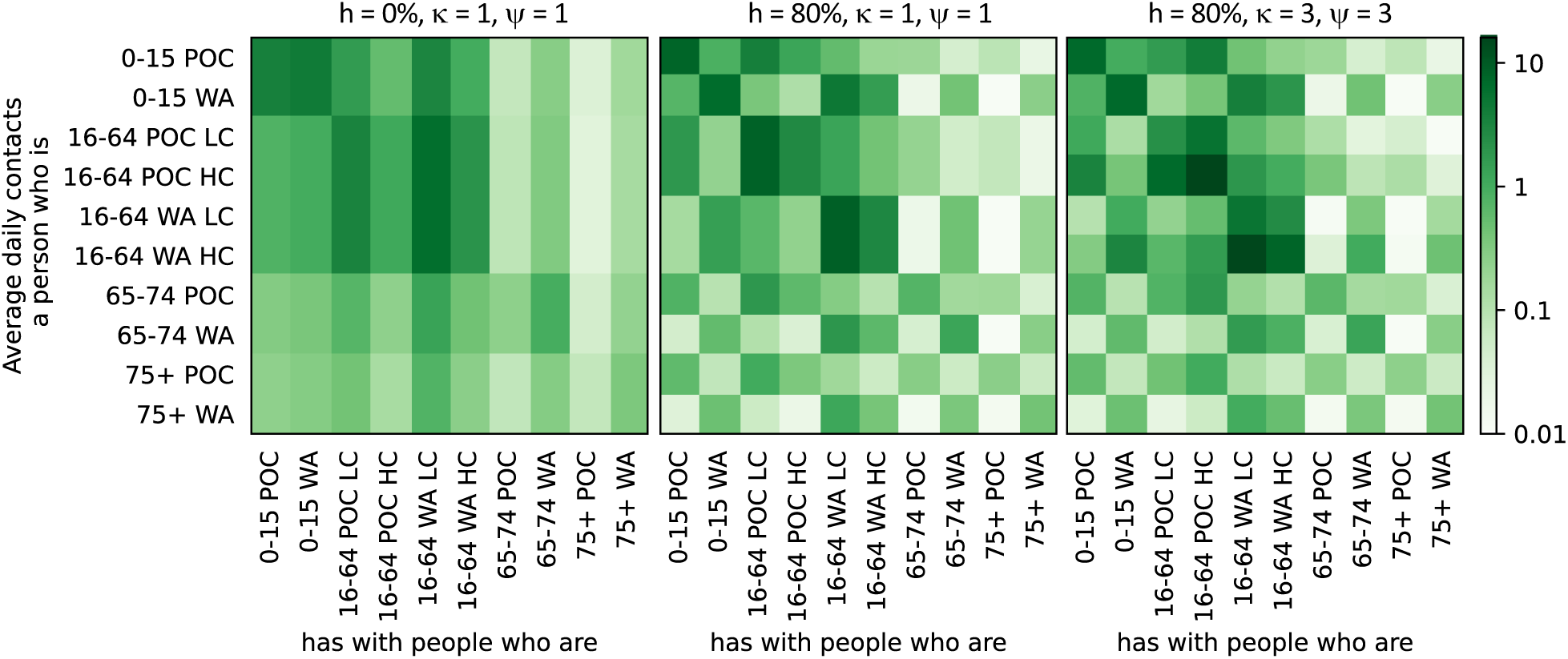
Contact patterns between sub-populations for different scenarios distinguished by the key parameters: *h, κ* and *ψ*.

### 2.3. Model dynamics

To model disease progression from infection to recovery or death, we distinguish for each sub-population 20 different stages as in Islam et al. (2021). At any moment, a person in sub-population *i* is either susceptible (*S*_*i*_), exposed and not yet infectious (*E*_*i*_), pre-symptomatic and infectious (*P*_*i*_), symptomatic and infectious (*C*_*i*_: clinical), asymptomatic and infectious at a lower rate (*A*_*i*_), quarantining and thus no longer infectious (*Q*_*i*_), recovered from symptomatic infection (*R*_*i*_), recovered from asymptomatic infection (*RA*_*i*_) or dead (*D*_*i*_). Note that reinfections were very rare prior to the emergence of the omicron variant (O Murchu et al., 2022; New York State Department of Health, 2022). As we study the spread of SARS-CoV-2 during the initial U.S. vaccine roll-out before the dominance of the omicron variant, we therefore do not model reinfections. We further distinguish individuals by their vaccination status. A person is either willing to get vaccinated and waiting for an available vaccine, not willing to get vaccinated, or already vaccinated, and as in Islam et al. (2021) we assume a vaccine hesitancy of 30% throughout the population.

The number of new infections in sub-population *i* at time *t* is given by *S*_*i*_(*t*)·Λ_*i*_(**P**(*t*), **C**(*t*), **A**(*t*)). Here, Λ_*i*_ is the *force of infection* function and includes, among other things, the above-defined contact matrix, which describes the mixing patterns (i.e., the average number of daily contacts) between all sub-populations. For other details, such as the entire system of ordinary differential equations describing the model dynamics as well as specific parameter choices, see Islam et al. (2021). Except for the contact matrix, we use all the same parameter choices. Note that we assume that any virus and disease parameters such as the case fatality rate or the rate of asymptomatic disease progression may depend on age but not on ethnicity. That is, differences between WA and POC are completely due to differences in the demographics and the contact matrix, which depends on the values of the three key parameters, *h, κ*, and *ψ*.

### 2.4. Vaccination and vaccine roll-out

Each vaccine allocation strategy assigns the ten sub-populations to a specific phase and may contain between one (i.e., all sub-populations vaccinated at the same time) and ten distinct phases. We follow the same approach as in Islam et al. (2021) to model daily vaccinations and vaccine function. In brief, we consider a population-wide, constant vaccine hesitancy rate of 30%, in line with estimates for early 2021, so that in each sub-population, 70% are willing to be vaccinated (Reiter et al., 2020). We use historic data on vaccinations to inform the daily number of newly vaccinated individuals, considering a single vaccination event (no two doses) for simplicity (CDC, 2021b). On any given day, if the number of available vaccines is less than the number of unvaccinated individuals willing to get vaccinated and part of the current phase, then we distribute the available doses equally (i.e., proportionally) among all these people. If there are more vaccines available than people waiting for the vaccine in the current phase, then everyone in the current phase gets vaccinated and the next phase of vaccinations begins.

Most people in the United States were vaccinated using the mRNA COVID-19 vaccines by Pfizer-BioNTech and Moderna. We used the same best estimates for three parameters related to vaccine function as in Islam et al. (2021), namely, vaccinated individuals are assumed to be 70% less likely to contract the virus, 66.7% less likely to develop a symptomatic infection when infected, and 50% less likely to transmit the virus (Pritchard et al., 2021). The first two parameters add up to a vaccine efficacy of 90% = 1 − (1 − 0.7)(1 − 0.667), as observed in the U.S. population in early 2021 (Thompson, 2021).

### 2.5. Model fitting

The previously developed model was fitted to cases and deaths observed between December 14, 2020 and April 29, 2021 assuming implementation of the vaccine allocation strategy suggested by the CDC (Islam et al., 2021). While we here consider a stratification of the U.S. population into different sub-populations, all parameters included in the model, as well as the total contact numbers and the total contacts between people of age group *i* and age group *j* are the same (see Contact patterns). We can therefore use the best fit parameter values from Islam et al. (2021).

### 2.6. Initial conditions

The U.S. vaccine roll-out began on December 14, 2020. To obtain the initial conditions (i.e., the number of individuals in each sub-population and each compartment on December 14, 2020), we match the model-predicted cases and deaths to the reported historic data from the New York Times COVID-19 database (New York Times, 2022). According to these data, the first case was reported on January 21, 2020. We therefore initialize the model with one symptomatic person (in compartment C), split between the different sub-populations relative to their sizes, and begin the simulations on *t*_0_ = January 21, 2020. For each sub-population, we further infer the number of people in the other infectious compartments on January 21, 2020 as

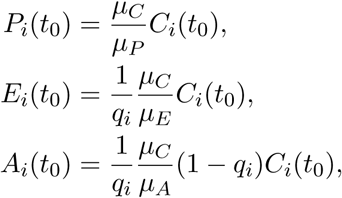

where 1*/µ*_*Y*_ is the average time spent in transient compartment *Y* and *q*_*i*_ is the probability of symptomatic infection (also known as clinical fraction). We further set *Q*_*i*_(*t*_0_) = *RA*_*i*_(*t*_0_) = *R*_*i*_(*t*_0_) = *D*_*i*_(*t*_0_) = 0 and *S*_*i*_(*t*_0_) = *N*_*i*_ − *E*_*i*_(*t*_0_) − *P*_*i*_(*t*_0_) − *C*_*i*_(*t*_0_) − *A*_*i*_(*t*_0_). We acknowledge that, due to under-reporting issues, the true numbers were likely higher than what is reported in the New York Times COVID-19 database, especially in early 2020.

We then run the model from January 21, 2020 until December 14, 2020. Each day, we record the number of new infections (i.e., *S*_*i*_ → *E*_*i*_) according to the model and re-scale as follows to match historic case numbers. Based on data released by the CDC, prior to the start of the vaccine roll-out, 10.27%, 75.65%, 7.70%, 6.38% of all reported cases were among people of age 0 − 15, 16 − 64, 65 − 74, 75+, respectively (CDC, 2021a). We assume an age-dependent rate of symptomatic infection *q*_*i*_ of 58.9%, 70%, 80.4% and 85% for the four age groups (Islam et al., 2021; CDC and ASPR, 2020). This implies that 10.70%, 78.82%, 5.87%, 4.61% of all infections occurred in the respective age group. For each age group and at each day, we re-scale the model-predicted new infections to fit the historic case data reported six days later, as the average time from infection to symptoms in our model is assumed to be six days (CDC and ASPR, 2020). That is, if we want to observe *x* (symptomatic) cases in age group *j* at day *t* + 6, we set the number of new infections in age group *j* at day *t* to *x/q*_*j*_. This procedure ensures an excellent fit between the model-predicted and historic case numbers, while the model determines the relative number of infected in each sub-population. To match historic deaths, we follow a similar procedure. Prior to the vaccine roll-out, 0.07%, 19.1%, 21.35% and 59.46% of all deaths had occurred in the age groups 0 − 15, 16 − 64, 65 − 74, 75+, respectively (CDC, 2021a). For each age group and at each day, we re-scale the model-predicted new deaths to fit the historic deaths for that day. We run the model from January 21, 2020 to December 14, 2020, and use the values from December 14, 2020 to initialize the population size of each compartment.

### 2.7. Outcome measures

Given a vaccine allocation strategy, we run the model from December 14, 2020 to December 31, 2021. Total predicted deaths by December 31, 2021 due to COVID-19 constitute the primary outcome measure used to evaluate vaccine allocation strategies. Secondary outcome measures are total predicted (symptomatic) cases by December 31, 2021, as well as predicted deaths and predicted cases among the WA and the POC sub-population. We focus primarily on total deaths as (i) this proved to be the primary vaccination goal in the U.S. public health response (National Academies of Sciences, Engineering, and Medicine, 2020) and (ii) it gives rise to more interesting trade-offs than e.g. minimizing total cases where only contact levels but not age-dependent case fatality rates need to be taken into account.

### 2.8. Global optimization approach

To find the best vaccine allocation strategy with respect to a given outcome measure, we perform an exhaustive evaluation of all possibly optimal strategies that assigned the ten sub-populations into 1, 2, 3, 4, 5, or 10 distinct phases. Any strategy that recommends vaccination of 16-64 year olds of a certain ethnicity group (WA or POC) with low-contact jobs before vaccinating people of the same age and ethnicity group with high-contact jobs can never be optimal since the opposite strategy would lead to lower incidence without increasing disease-related mortality (on average, all 16-64 year olds have the same case fatality rate). We therefore exclude all such strategies from the search. For 1, 2, 3, 4, 5, or 10 distinct phases there are 1, 574, 24519, 308076, 1724310 and 907200 possibly optimal strategies, respectively. For each strategy, we run the model starting on December 14, 2020, the beginning of the U.S. vaccine roll-out, until December 31, 2021 and record all outcome measures (see above). We do not investigate strategies with 6-9 distinct phases because (i) as we will show, the improvement in outcome measures going from 4 to 5 phases is very small, and (ii) many distinct phases are hard to implement from a public health perspective.

### 2.9. Scenario analysis

We conducted global optimizations for six scenarios that differ in their choices for the three key model parameters (2 choices each): 1. ethnic homophily *h*, 2. relative contact level of people employed in high-contact jobs compared to low-contact jobs (*κ*), 3. relative proportion of POC compared to WA who work high-contact jobs (*ψ*). We considered a baseline value of *h* = 0% (i.e., no homophily) and *h* = 80% (high homophily). The former value is used in all infectious disease models that do not explicitly account for homophily, while the latter is likely a more realistic estimate (Mollica et al., 2003). For the other two key model parameters, we considered a baseline value of 1 (i.e., no difference) and 3 (i.e., a three-fold increased number of contacts by employees in high-contact jobs, or a three-fold increased proportion of POC in high-contact jobs). We considered all combinations of parameter choices, except those two with *κ* = 1, *ψ* = 3 as the choice of *ψ* is irrelevant if *κ* = 1 (i.e., if *κ* = 1, there is no difference between high- and low-contact jobs). The two main scenarios we deem important are parametrized by

1. *h* = 0%, *κ* = 1, *ψ* = 1: the base scenario. These assumptions are intrinsically used in any model that does not account for ethnic homophily nor different occupation-mediated average contact levels for POC and WA.
2. *h* = 80%, *κ* = 3, *ψ* = 3: the most likely social context. While we lack reliable estimates for all three parameters, it is clear that *h >>* 0, *κ >* 1, *ψ >* 1 (Mollica et al., 2003; US Bureau of Labor Statistics, 2021a,b).

## 3. Results and Discussion

In December 2020, the CDC recommended structuring the U.S. COVID-19 vaccine roll-out in four phases and prioritized vaccination of older people (65+) in congested living conditions as well as healthcare workers and frontline essential workers with many contacts (Dooling et al., 2020). The CDC strategy did not differentiate between ethnic groups. We find here that allowing for differences in occupational risk level but no ethnic homophily (*h* = 0%, *κ* = 3), the 4-phase allocation strategy that minimizes mortality is to first vaccinate people 75 and older, then vaccinate people employed in high-contact jobs (e.g., healthcare and frontline essential workers), then people 65-74 years old, followed by the remainder of the population (Table 1). This is consistent with the strategy suggested by the CDC. Note that in a previous model, we showed that the only way to improve the CDC strategy was to differentiate between people with and without known comorbidities (Islam et al., 2021).

**Table 1:**
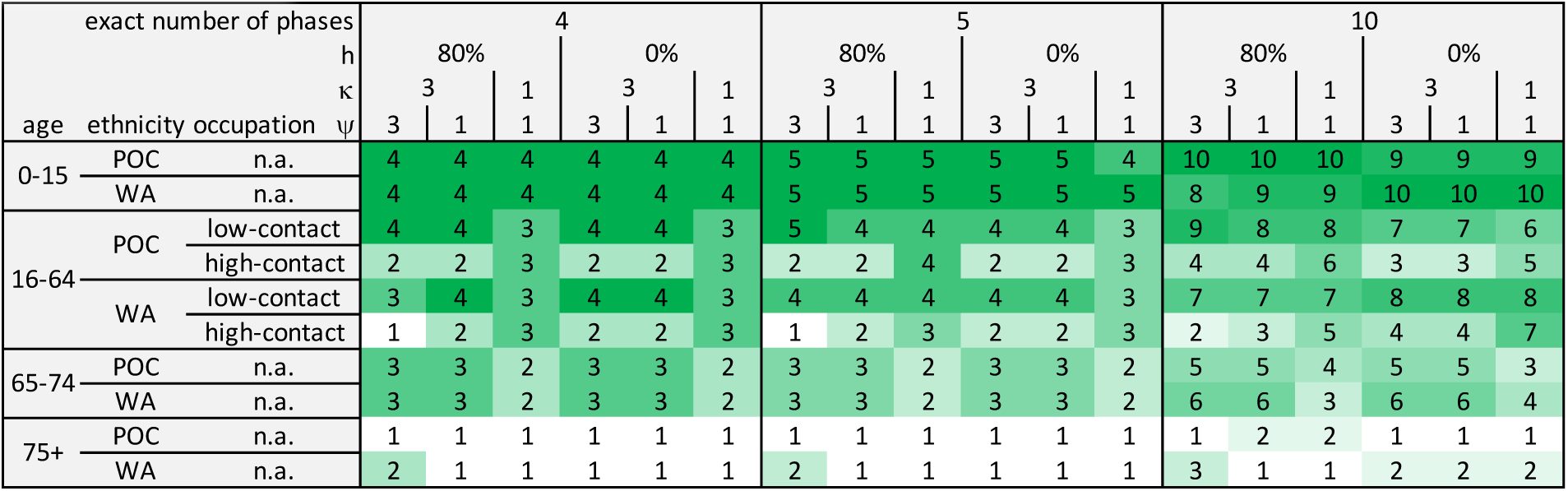
Optimal vaccine allocation strategies to minimize COVID-19 mortality given a different number of phases and for different scenarios specified by the level of ethnic homophily (*h*), relative contact levels for employees in high-contact jobs (compared to low-contact jobs) (*κ*), and the relative proportion of POC (compared to WA) in high-contact jobs (*ψ*).

### 3.1. Number of vaccine roll-out phases

Notwithstanding the increased logistical complexity entailed in a roll-out with many phases, we investigated the effect of changing the number of phases on the primary outcome metric, total COVID-19 deaths. For each scenario, we compared the predicted deaths from the best strategy with a given number of phases to the overall best strategy with any number of phases (1-5 or 10). More phases provide more opportunity for fine-tuning, and it was thus not surprising that a vaccine roll-out without priority phases (i.e., with every sub-population in the same single phase) led to the highest predicted death count (Table 2). The introduction of more phases generally led to fewer deaths, but the marginal gain per added phase decreased rapidly.

**Table 2:**
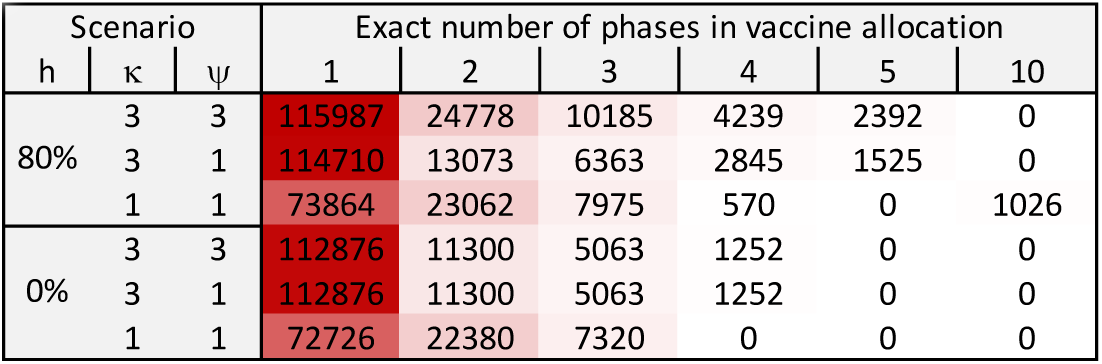
For each scenario (rows), we identified the overall best allocation strategy in minimizing deaths (no matter the number of phases) and computed excess predicted deaths when using the optimal allocation strategy with exactly *n* = 1, 2, 3, 4, 5, 10 phases.

The more complex a scenario, the more phases were needed to achieve the best outcome. In the base scenario (*h* = 0%, *κ* = 1, *ψ* = 1, corresponding to no ethnic homophily and no differences in job-related contact levels), four phases sufficed to achieve the lowest possible number of deaths because this scenario only stratifies the population by age; the optimal 4-phase allocation was to prioritize vaccine access strictly by age (Table 1). When considering ethnic homophily (80%) but no differences in occupation (*κ* = 1, *ψ* = 1), an allocation with five phases led to a slightly better predicted outcome than any 4-phase allocation. This is because the optimal 5-phase allocation allows for structuring the vaccine roll-out among the 16-64 year olds in two distinct phases, enabling prioritization of a subset of WA since this ethnicity group contains relatively more at-risk elderly people (see Figure 1a). Finally, in the scenario resembling the most likely social context (*h* = 80%, *κ* = 3, *ψ* = 3), the optimal 10-phase allocation strategy was better than any strategy with five or fewer phases.

For one scenario (*h* = 80%, *κ* = 1, *ψ* = 1), all 10-phase allocation strategies led to more deaths than the optimal 5-phase strategy. This scenario does not distinguish between high- and low-contact jobs; both types of jobs have the same number of contacts (*κ* = 1), and WA and POC are equally engaged in both types of jobs (*ψ* = 1). Given the presence of ethnic homophily (*h* = 80%), it is optimal to vaccinate both WA and POC at the same time and rate, which is possible using exactly five phases but not when forcing ten phases (see optimal 5-phase and 10-phase allocations for this scenario in Table 1).

### 3.2. Trade-offs in vaccine prioritization

The fundamental trade-off in COVID-19 vaccine prioritizations centers on age. Should we protect the most vulnerable people directly by vaccinating them first, or indirectly by vaccinating the most socially active people first, thereby achieving the best possible incidence reduction and lowering the risk of infection for the vulnerable population? The timing of the vaccine roll-out certainly affects this trade-off. In a time of low incidence, the indirect option may be suitable, while at the peak of an epidemic wave it may be too costly to wait for the indirect effect to manifest. We only derived optimal allocation strategies for the specific timing of the U.S. vaccine roll-out, which commenced on December 14, 2020, during a time of high incidence. Given this, the optimal vaccine allocation strategy in the simplest base scenario was to prioritize vaccine access solely by age (Table 1). This is because older people, despite their lower average contact levels (e.g., 16-64 year olds have 5.1 times more contacts than people 75 and older), have a dramatically higher case fatality rate (e.g., the CFR of people 75 and older is 37x higher than for 16-64 year olds and even 1300x higher than for children) (Figure 1a).

When considering the most likely social context (*h* = 80%, *κ* = 3, *ψ* = 3), this simple reasoning no longer works. First, the presence of homophily means that vaccination of one ethnicity group (POC or WA) mainly provides indirect protection to individuals from the same ethnicity group. Second, WA are on average older, so the indirect protection may be more important for WA than for POC, where relatively fewer contacts occur between old (65+) and young. Third, POC work more high-contact jobs, so the incidence among POC, in the absence of any vaccines, will likely be higher. The second point suggests WA should be prioritized over POC, while the third point suggests the opposite, illustrating the second trade-off in vaccine prioritization: the prioritization between ethnicity groups, which we investigate in this study.

These considerations help explain the patterns observed in the optimal 4-, 5- and 10-phase allocation strategies for this scenario (Table 1). As expected, all three optimal strategies exhibit the same relative order, up to ties in the 4- and 5-phase strategies. In all three optimal strategies (when given the choice by allowing for a sufficient number of phases), older POC (65+) are vaccinated before WA of the same age group, while younger WA are vaccinated before POC of the same age group and job type. Interestingly, vaccinating 75+ year old individuals before 16-64 year old employees in high-contact jobs is ideal for POC, whereas the opposite is the case for WA. This is an instance where the high incidence rates among POC (due to many employees in high-contact jobs) favor direct protection of the most vulnerable POC. For WA, on the other hand, indirect protection proved favorable, (i) because relatively more contacts occur between old (65+) and young WA (compared to POC with a smaller proportion of people 65 and older), and (ii) due to age assortativity in contact matrices: the oldest POC are already vaccinated, providing—despite high homophily—a reduction in infections of 75+ year old WA by 75+ year old POC.

### 3.3. Accounting for ethnicity in vaccine prioritization

As shown above, taking ethnicity into account when prioritizing vaccine access may reduce COVID-19 deaths. To investigate this further, we compared, for each scenario and two to five phases, the predicted mortality under the respective optimal allocation strategy and under the respective optimal strategy that does not differentiate by ethnicity (that is, which vaccinates all people of same age and occupation at the same time; Table 3). For the basic scenarios which assume no ethnic homophily (*h* = 0%), ethnicity does not need to be taken into consideration to achieve the lowest possible death count. However, in the more likely social context (*h* = 80%) and especially when also assuming proportionately more POC in high-contact jobs (*κ* = 3, *ψ* = 3), vaccine roll-outs that do not differentiate by ethnicity lead to more predicted deaths. The gap was larger for allocation strategies with several phases. This finding clearly highlights the importance of understanding the impact of ethnicity and the related parameters on vaccine roll-out design, and more generally, on disease dynamics.

**Table 3:**
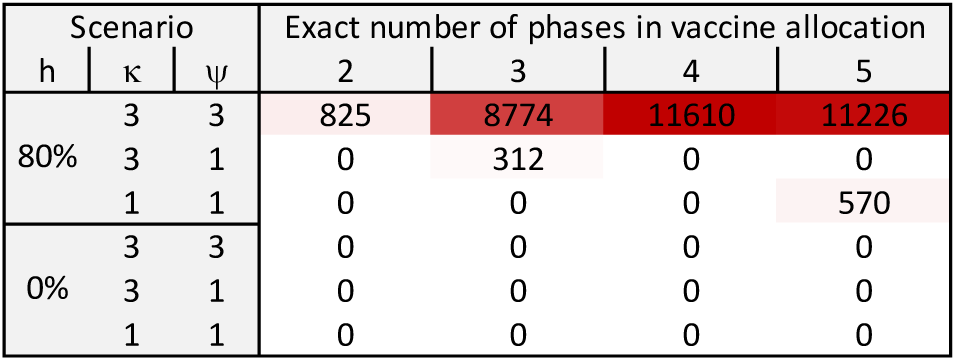
Excess predicted deaths when not differentiating vaccine access by ethnicity. For all scenarios (rows) and 2-5 vaccine allocation phases (columns), the predicted deaths under the respective best strategy are compared to those under the best strategy that assigns WA and POC with same characteristics (age and occupation) into the same phase.

| Scenario |  |  | Exact number of phases in vaccine allocation |  |  |  |
| --- | --- | --- | --- | --- | --- | --- |
| $h$ | $\kappa$ | $\psi$ | 2 | 3 | 4 | 5 |
| 80% | 3 | 3 | 825 | 8774 | 11610 | 11226 |
|  | 3 | 1 | 0 | 312 | 0 | 0 |
|  | 1 | 1 | 0 | 0 | 0 | 570 |
| 0% | 3 | 3 | 0 | 0 | 0 | 0 |
|  | 3 | 1 | 0 | 0 | 0 | 0 |
|  | 1 | 1 | 0 | 0 | 0 | 0 |

### 3.4. Trade-offs among vaccination goals

Thus far, we have focused solely on reducing total COVID-19 deaths. Other possible vaccination goals include reducing the total number of cases, and reducing deaths or cases in a specific ethnic group (e.g., because POC are disproportionately affected by COVID-19 (Garcia et al., 2021; Labgold et al., 2021)). All these goals conflict with each other. A pairwise Spearman correlation analysis over all possibly optimal 10-phase vaccine allocation strategies quantifies the extent of these trade-offs (Figure 3). Strategies that are good at reducing overall deaths are also good at reducing deaths among WA people. The same is true to a lesser extent for cases, and likely due to the predominance of WA in the U.S. population and especially among the elderly, most at-risk population (Figure 1a). Furthermore, the correlations between the vaccination goals differ strongly between the base scenario and the most likely social context. When considering the correlations over all possibly optimal strategies with four or five phases instead, the differences were very small (not shown).

**Figure 3:**
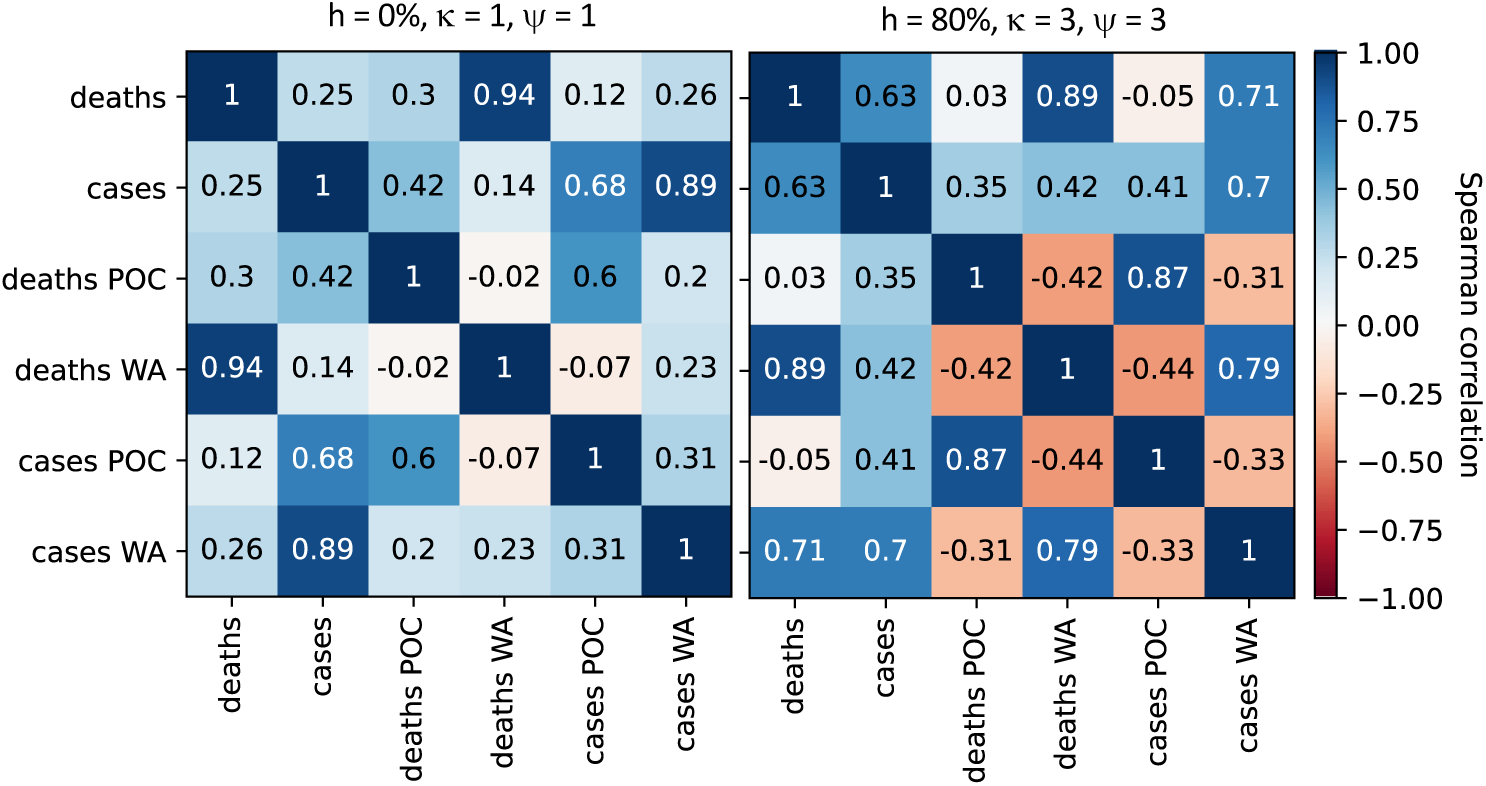
Pairwise Spearman correlation for six different outcome metrics (i.e., possible vaccination goals), for the base scenario (*h* = 0%, *κ* = 1, *ψ* = 1; left) and the most likely social context (*h* = 80%, *κ* = 3, *ψ* = 3; right). The correlations are taken over the outcomes from all possibly optimal 10-phase vaccine allocation strategies. Positive (negative) correlations are shown in blue (red) and the color intensity describes the absolute correlation strength.

The correlation analysis shows that it is generally impossible to design a strategy that is simultaneously optimal in multiple vaccination objectives. The concept of Pareto-optimality can be used to describe strategies that are “optimal” in multiple objectives. A strategy is Pareto-optimal for a given set of objectives if there is no strategy that performs better in one objective without performing worse in another. An analysis of the Pareto frontier of all Pareto-optimal 10-phase strategies for three pairs of opposing vaccination goals reveals several interesting results. First, strategies that only focus on reducing deaths among one of the two ethnicity groups lead to non-Pareto-optimal outcomes in terms of total deaths and cases (Figure 4a,d). Especially in the most likely social context (*h* = 80%, *κ* = 3, *ψ* = 3), the number of both deaths and cases is substantially higher (Table 4). Second, the optimal strategy reducing total cases is more similar to the optimal strategy reducing cases among WA (Kendall’s *τ* coefficient = 0.56 between the two allocations) than to the optimal strategy reducing cases among POC (*τ* = 0.24; Table 4). This is probably because 65.8% of the U.S. population is WA. The predominance of WA people likely also explains why the optimal strategy reducing total cases always prioritizes vaccination of WA before POC of the same age group and the same job type, and even suggests vaccinating WA 65-74 year olds with fewer contacts before vaccinating POC children (Table 4).

**Figure 4:**
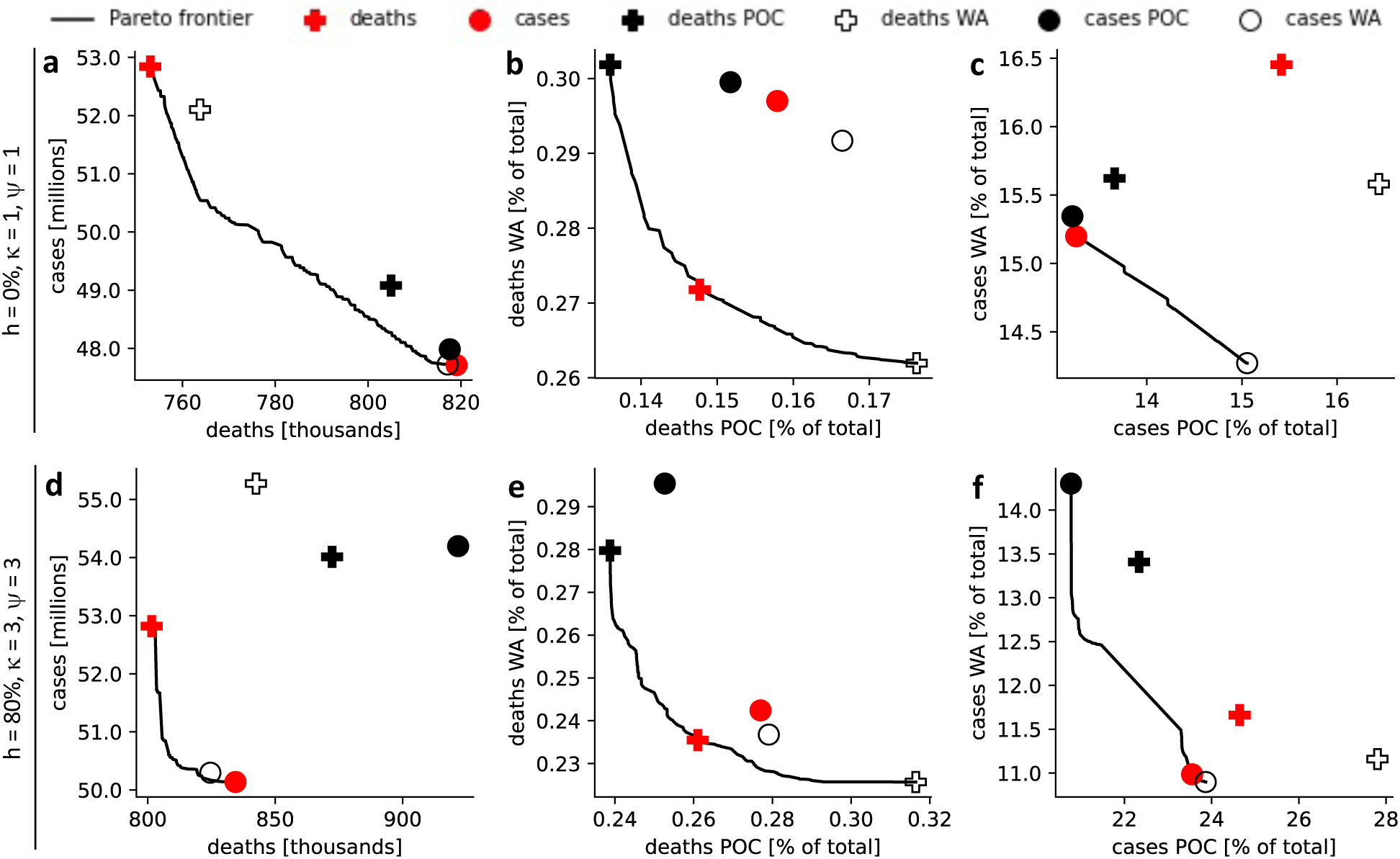
Trade-offs between vaccination goals. The performance in multiple objectives (total deaths and cases, as well as ethnicity group-specific deaths and cases) is shown for each of the 10-phase vaccine allocations, which are optimal in one of the objectives. 2-dimensional Pareto frontiers display the performance of all 10-phase vaccine allocations, which cannot be improved in one objective without a worse outcome in the other objective shown in the sub panel. (a-c) base scenario with *h* = 0%, *κ* = 1, *ψ* = 1, (d-f) most likely social context with *h* = 80%, *κ* = 3, *ψ* = 3. (a,d) total deaths and total cases, (b,e) proportion of all POC and WA who died from COVID-19, (c,f) proportion of all POC and WA with a symptomatic COVID-19 infection.

**Table 4:** Optimal 10-phase vaccine allocation strategy (shaded in green) for six different objectives (columns), for the scenario corresponding to the most likely social context (*h* = 80%, *κ* = 3, *ψ* = 3). For each strategy, the absolute and relative performance in all six objectives is shown below (shaded in red based on the relative performance). Absolute numbers: deaths in thousands, cases in millions.

| sub-population characteristics |  |  | optimal 10-phase vaccine allocation strategy minimizing |  |  |  |  |  |
| --- | --- | --- | --- | --- | --- | --- | --- | --- |
| age | ethnicity | occupation | deaths | cases | deaths POC | deaths WA | cases POC | cases WA |
| 0-15 | POC | n.a. | 10 | 7 | 6 | 7 | 6 | 7 |
|  | WA | n.a. | 8 | 5 | 8 | 5 | 7 | 5 |
| 16-64 | POC | low-contact | 9 | 4 | 5 | 10 | 2 | 10 |
|  |  | high-contact | 4 | 2 | 3 | 6 | 1 | 2 |
|  | WA | low-contact | 7 | 3 | 7 | 4 | 8 | 3 |
|  |  | high-contact | 2 | 1 | 4 | 1 | 3 | 1 |
| 65-74 | POC | n.a. | 5 | 8 | 2 | 9 | 4 | 9 |
|  | WA | n.a. | 6 | 6 | 10 | 3 | 10 | 4 |
| 75+ | POC | n.a. | 1 | 10 | 1 | 8 | 5 | 8 |
|  | WA | n.a. | 3 | 9 | 9 | 2 | 9 | 6 |
| Respective outcome of specific allocation | deaths |  | 801.6 | 834.4 | 872.3 | 842.5 | 921.7 | 824.6 |
|  | cases |  | 52.82 | 50.14 | 54.01 | 55.27 | 54.20 | 50.30 |
|  | deaths POC |  | 292.4 | 310.3 | 267.5 | 354.5 | 283.0 | 312.7 |
|  | deaths WA |  | 509.1 | 524.1 | 604.9 | 488.0 | 638.7 | 511.9 |
|  | cases POC |  | 27.61 | 26.38 | 25.02 | 31.15 | 23.27 | 26.73 |
|  | cases WA |  | 25.21 | 23.75 | 28.99 | 24.13 | 30.92 | 23.56 |
| Difference in outcome between specific and respective best allocation | deaths |  | 0% | 4.1% | 8.8% | 5.1% | 15.0% | 2.9% |
|  | cases |  | 5.4% | 0% | 7.7% | 10.2% | 8.1% | 0.3% |
|  | deaths POC |  | 9.3% | 16.0% | 0% | 32.5% | 5.8% | 16.9% |
|  | deaths WA |  | 4.3% | 7.4% | 24.0% | 0% | 30.9% | 4.9% |
|  | cases POC |  | 18.6% | 13.4% | 7.5% | 33.8% | 0% | 14.9% |
|  | cases WA |  | 7.0% | 0.8% | 23.0% | 2.4% | 31.2% | 0% |

In the base scenario, the per-capita death rate for WA is substantially higher than for POC, even when following an optimal vaccine allocation which only minimizes deaths among WA (Figure 4b). In the most likely social context (*h* = 80%, *κ* = 3, *ψ* = 3), however, it is very similar between the two ethnicity groups (Figure 4e), which aligns closely with data from late 2021 (Kaiser Family Foundation, 2022). This provides further evidence that this scenario likely describes the real social context better than the base scenario. Note that a similar per-capita death rate does not imply POC and WA are equally affected by COVID-19. When adjusted for the lower average age of POC, substantially fewer deaths per POC would be expected (CDC, 2022; Gold et al., 2020). The increased per-capita rate of symptomatic infection also shows the disproportionate burden of COVID-19 on POC (Figure 4f) and aligns with reported data (CDC, 2022; Kaiser Family Foundation, 2022).

## 4. Limitations and Conclusions

Our study has several limitations. First, we only consider two ethnicity groups, POC and WA. We made this choice to (i) have a manageable number of sub-populations allowing us to use the global optimization approach, and (ii) be able to describe the level of homophily using a single parameter. In reality, the U.S. society has many ethnic groups, and the rate of mixing between any two ethnic groups may differ. Second, we assume all disease parameters for POC and WA are the same, and there are no differences due to e.g. genetic factors. However, a higher prevalence of COVID-19 risk factors has been reported for POC, resulting in higher case fatality rates for POC (Kabarriti et al., 2020). When included, this could alter the model outcomes but would also complicate the interpretation of the results, which in this study only depend on the three key model parameters and demographic differences. Third, the specific choice of optimal allocation strategy may be sensitive to the timing of the vaccine roll-out (e.g., the incidence level and the level of immunity due to natural infection in one ethnic group versus another). Elucidating this impact of timing would be an interesting avenue for future, more theoretical studies.

While accurate data on social contact patterns is difficult to collect (Mossong et al., 2008; Prem et al., 2017), this study highlights the importance of ethnic homophily on disease dynamics, and more specifically, on the design of optimal mass vaccine roll-out strategies. The most likely social context in the U.S. is very different from standard model assumptions which do not account for ethnicity and ethnic homophily, and this difference significantly affects which vaccination strategy is optimal. It may thus be worth exploring options to better quantify social contact patterns between ethnic groups to achieve a better understanding of infectious disease spread.

## Data Availability

All data produced in the present work are contained in the manuscript, all relevant code is available at GitHub: https://github.com/ckadelka/Ethnicity-and-vaccine-prioritization.

## CRediT authorship contribution statement

**Claus Kadelka**: Conceptualization, Methodology, Software, Formal analysis, Writing - Original Draft, Writing - Review & Editing, Visualization, Supervision, **Md Rafiul Islam**: Conceptualization, Methodology, Writing - Review & Editing, Supervision, **Audrey McCombs**: Methodology, Writing - Original Draft, Writing - Review & Editing, **Jake Alston**: Methodology, Writing - Review & Editing, **Noah Morton**: Methodology, Writing - Review & Editing

## Declaration of competing interest

The authors declare that they have no known competing financial interests or personal relationships that could have appeared to influence the work reported in this paper.

## Acknowledgments

The authors received no funding for this study. This study began as a semester-long undergraduate research project. We thank the undergraduate researchers Emma Staut, Caroly Coronado-Vargas, Kassandra Chino Gonzalez, and Benjamin Studebaker for fruitful discussions in the early stage of this project. We also thank Preeti Sar who served as a mentor for the students.

## Data availability

Data and all relevant code is available at GitHub: https://github.com/ckadelka/Ethnicity-and-vaccine-prioritization.

## Notes

### Competing Interest Statement

The authors have declared no competing interest.

### Funding Statement

This study did not receive any funding

## References

Bhala N, Curry G, Martineau AR, Agyemang C, Bhopal R (2020) Sharpening the global focus on ethnicity and race in the time of COVID-19. The Lancet 395(10238):1673–1676

Bubar KM, Reinholt K, Kissler SM, Lipsitch M, Cobey S, Grad YH, Larremore DB (2021) Model-informed COVID-19 vaccine prioritization strategies by age and serostatus. Science 371(6532):916–921

Burgio G, Steinegger B, Arenas A (2022) Homophily impacts the success of vaccine roll-outs. Communications Physics 5(1):1–7

CDC (2020) Introduction to COVID-19 racial and ethnic health disparities. https://www.cdc.gov/coronavirus/2019-ncov/community/health-equity/racial-ethnic-disparities/index.html (version July 1, 2022)

CDC (2021a) COVID-19 case surveillance public use data. https://data.cdc.gov/Case-Surveillance/COVID-19-Case-Surveillance-Public-Use-Data-with-Ge/n8mc-b4w4

CDC (2021b) Trends in number of COVID-19 vaccinations in the US. https://covid.cdc.gov/covid-data-tracker/#vaccination-trends

CDC (2022) Risk for COVID-19 infection, hospitalization, and death by race/ethnicity. https://www.cdc.gov/coronavirus/2019-ncov/covid-data/investigations-discovery/hospitalization-death-by-race-ethnicity.html (version June 2, 2022)

CDC and ASPR (2020) COVID-19 pandemic planning scenarios by the CDC and the Office of the Assistant Secretary for preparedness and response (ASPR). https://www.cdc.gov/coronavirus/2019-ncov/hcp/planning-scenarios.html (version Mar 19, 2021)

Cox D, Navarro-Rivera J, Jones RP, et al. (2016) Race, religion, and political affiliation of americans’ core social networks. Public Religion Research Institute

Currarini S, Jackson MO, Pin P (2009) An economic model of friendship: Homophily, minorities, and segregation. Econometrica 77(4):1003–1045

Dooling K, Marin M, Wallace M, McClung N, Chamberland M, Lee GM, Talbot HK, Romero JR, Bell BP, Oliver SE (2020) The advisory committee on immunization practices’ interim recommendation for allocating initial supplies of COVID-19 Vaccine — United States, 2020. Morbidity and Mortality Weekly Report 69(49):1857–1859

Feng Z, Hill AN, Smith PJ, Glasser JW (2015) An elaboration of theory about preventing outbreaks in homogeneous populations to include heterogeneity or preferential mixing. Journal of theoretical biology 386:177–187

Foy BH, Wahl B, Mehta K, Shet A, Menon GI, Britto C (2021) Comparing covid-19 vaccine allocation strategies in india: A mathematical modelling study. International Journal of Infectious Diseases 103:431–438

Funk S, Knapp JK, Lebo E, Reef SE, Dabbagh AJ, Kretsinger K, Jit M, Edmunds WJ, Strebel PM (2019) Combining serological and contact data to derive target immunity levels for achieving and maintaining measles elimination. BMC medicine 17(1):1–12

Garcia MA, Homan PA, García C, Brown TH (2021) The color of covid-19: Structural racism and the disproportionate impact of the pandemic on older black and latinx adults. The Journals of Gerontology: Series B 76(3):e75–e80

Gold JA, Rossen LM, Ahmad FB, Sutton P, Li Z, Salvatore PP, Coyle JP, DeCuir J, Baack BN, Durant TM, et al. (2020) Race, ethnicity, and age trends in persons who died from COVID-19—United States, May–August 2020. Morbidity and Mortality Weekly Report 69(42):1517

Hiraoka T, Rizi AK, Kivelä M, Saramäki J (2022) Herd immunity and epidemic size in networks with vaccination homophily. Physical Review E 105(5):L052,301

Hooper MW, Nápoles AM, Pérez-Stable EJ (2020) COVID-19 and racial/ethnic disparities. JAMA 323(24):2466–2467

Ingraham C (2014) Three quarters of whites don’t have any non-white friends. Washington Post 25, URL https://www.washingtonpost.com/news/wonk/wp/2014/08/25/three-quarters-of-whites-dont-have-any-non-white-friends/

Islam MR, Oraby T, McCombs A, Chowdhury MM, Al-Mamun M, Tyshenko MG, Kadelka C (2021) Evaluation of the United States COVID-19 vaccine allocation strategy. PloS one 16(11):e0259.700

Kabarriti R, Brodin NP, Maron MI, Guha C, Kalnicki S, Garg MK, Racine AD (2020) Association of race and ethnicity with comorbidities and survival among patients with COVID-19 at an urban medical center in New York. JAMA network open 3(9):e2019.795–e2019,795

Kadelka C (2022) Projecting social contact matrices to populations stratified by binary attributes with known homophily. arXiv

Kadelka C, McCombs A (2021) Effect of homophily and correlation of beliefs on COVID-19 and general infectious disease outbreaks. PloS one 16(12):e0260.973

Kaiser Family Foundation (2022) COVID-19 cases and deaths by race/ethnicity: Current data and changes over time. URL https://www.kff.org/coronavirus-covid-19/issue-brief/covid-19-cases-and-deaths-by-race-ethnicity-current-data-and-changes-over-time/

Khanijahani A, Iezadi S, Gholipour K, Azami-Aghdash S, Naghibi D (2021) A systematic review of racial/ethnic and socioeconomic disparities in COVID-19. International journal for equity in health 20(1):1–30

Labgold K, Hamid S, Shah S, Gandhi NR, Chamberlain A, Khan F, Khan S, Smith S, Williams S, Lash TL, et al. (2021) Estimating the unknown: greater racial and ethnic disparities in COVID-19 burden after accounting for missing race/ethnicity data. Epidemiology (Cambridge, Mass) 32(2):157

März JW, Molnar A, Holm S, Schlander M (2022) The ethics of COVID-19 vaccine allocation: Don’t forget the trade-offs! Public Health Ethics

Matrajt L, Eaton J, Leung T, Brown ER (2021a) Vaccine optimization for COVID-19: Who to vaccinate first? Science Advances 7:eabf1374. DOI 10.1126/sciadv.abf1374

Matrajt L, Eaton J, Leung T, Dimitrov D, Schiffer JT, Swan DA, Janes H (2021b) Optimizing vaccine allocation for COVID-19 vaccines shows the potential role of single-dose vaccination. Nature Communications 12(1):1–18

McDonald CJ, Baik SH, Zheng Z, Amos L (2020) A method for prioritizing risk groups for early SARS-CoV-2 vaccination, by the numbers. medRxiv

McPherson M, Smith-Lovin L, Cook JM (2001) Birds of a feather: Homophily in social networks. Annual review of sociology pp 415–444

Millett GA, Jones AT, Benkeser D, Baral S, Mercer L, Beyrer C, Honermann B, Lankiewicz E, Mena L, Crowley JS, et al. (2020) Assessing differential impacts of COVID-19 on black communities. Annals of epidemiology 47:37–44

Mollica KA, Gray B, Trevino LK (2003) Racial homophily and its persistence in newcomers’ social networks. Organization science 14(2):123–136

Mossong J, Hens N, Jit M, Beutels P, Auranen K, Mikolajczyk R, Massari M, Salmaso S, Tomba GS, Wallinga J, et al. (2008) Social contacts and mixing patterns relevant to the spread of infectious diseases. PLoS medicine 5(3):e74

National Academies of Sciences, Engineering, and Medicine (2020) Framework for equitable allocation of COVID-19 vaccine. National Academies Press Washington, DC, https://doi.org/10.17226/25917

New York State Department of Health (2022) New York State statewide COVID-19 reinfection data. Data retrieved from https://health.data.ny.gov/Health/New-York-State-Statewide-COVID-19-Reinfection-Data/7aaj-cdtu, updated June 22, 2022

New York Times (2022) Coronavirus (COVID-19) data in the United States. https://github.com/nytimes/covid-19-data

O Murchu E, Byrne P, Carty PG, De Gascun C, Keogan M, O’Neill M, Harrington P, Ryan M (2022) Quantifying the risk of SARS-CoV-2 reinfection over time. Reviews in medical virology 32(1):e2260

Prem K, Cook AR, Jit M (2017) Projecting social contact matrices in 152 countries using contact surveys and demographic data. PLoS computational biology 13(9):e1005.697

Pritchard E, Matthews PC, Stoesser N, Eyre DW, Gethings O, Vihta KD, Jones J, House T, VanSteenHouse H, Bell I, et al. (2021) Impact of vaccination on new SARS-CoV-2 infections in the United Kingdom. Nature medicine 27(8):1370–1378

Reiter PL, Pennell ML, Katz ML (2020) Acceptability of a COVID-19 vaccine among adults in the United States: How many people would get vaccinated? Vaccine 38(42):6500–6507

Salathé M, Bonhoeffer S (2008) The effect of opinion clustering on disease outbreaks. Journal of The Royal Society Interface 5(29):1505–1508

Shadmi E, Chen Y, Dourado I, Faran-Perach I, Furler J, Hangoma P, Hanvoravongchai P, Obando C, Petrosyan V, Rao KD, et al. (2020) Health equity and COVID-19: global perspectives. International journal for equity in health 19(1):1–16

Tai DBG, Shah A, Doubeni CA, Sia IG, Wieland ML (2021) The disproportionate impact of COVID-19 on racial and ethnic minorities in the United States. Clinical Infectious Diseases 72(4):703–706

Thompson MG (2021) Interim estimates of vaccine effectiveness of BNT162b2 and mRNA-1273 COVID-19 vaccines in preventing SARS-CoV-2 infection among health care personnel, first responders, and other essential and frontline workers—eight US locations, December 2020–March 2021. Morbidity and Mortality Weekly Report 70

US Bureau of Labor Statistics (2021a) Employed persons by detailed occupation, sex, race, and Hispanic or Latino ethnicity. URL https://www.bls.gov/cps/cpsaat11.htm

US Bureau of Labor Statistics (2021b) Labor force characteristics by race and ethnicity, 2020. Bureau of Labor Statistics Report 1095, URL https://www.bls.gov/opub/reports/race-and-ethnicity/2020/home.htm

US Census Bureau (2020) Annual estimates of the resident population by sex, age, race, and Hispanic origin for the United States: April 1, 2010 to july 1, 2019. US Census Bureau Population Division URL https://www.census.gov/newsroom/press-kits/2020/population-estimates-detailed.html

